# Identifying candidate items for a prototype index on propensity to integrate research evidence into clinical decision-making in rehabilitation

**DOI:** 10.1101/2024.07.16.24310286

**Authors:** Jacqueline Roberge-Dao, Aliki Thomas, Annie Rochette, Keiko Shikako, Nancy Mayo

## Abstract

**Purpose:** Existing measures of evidence-based practice (EBP) in rehabilitation provide a fragmented interpretation of EBP competencies as performance is rated on discrete domains without a harmonized measure to represent the multidimensionality of EBP. Building on previous work, this study aimed to provide evidence that a brief multidimensional index can be formed to determine a clinician’s propensity to integrate research evidence into decision-making to inform the subsequent development process.

**Methods:** Using a Canadian dataset of occupational and physical therapists (n=127) who responded to a survey containing 70 frequently used items to measure EBP (representing six domains), one item per key EBP domain was selected using Rasch measurement theory and expert consensus. A preliminary scoring algorithm was developed for testing purposes. The interpretability of the prototype index was examined across characteristics of the sample and compared to full EBP measures using generalized estimating equations.

**Results:** Five items were selected for inclusion in the prototype index representing the dimensions of *use of research evidence, self-efficacy, resources, attitudes,* and *activities*. Testing demonstrated that the prototype behaves consistently regardless of age groups, gender, and setting, and provides comparable information to full EBP measures.

**Conclusion:** This study provides promising preliminary evidence to justify continuing the index development process. The benefits of having a global index of EBP as opposed to having multiple domain-specific measures are discussed in this paper.

## Introduction

With the aim of optimizing health and quality of life, rehabilitation clinicians must draw on various sources of knowledge to make decisions related to the care of patients. One approach that has been highly advocated in the last three decades for informing clinical decision-making (CDM) and improving the quality of clinical practice is evidence-based practice (EBP). Occupational therapists (OTs) and physical therapists (PTs) are expected to engage in EBP which involves integrating the best available research evidence, clinical expertise and patient preferences within the context of available resources.^1–3^

Despite over 30 years of developments in the application and study of EBP, the measurement of EBP is rife with challenges. Measurement of EBP is essential to identify clinicians’ level of engagement in EBP and the personal or organizational factors that should be improved or maintained. More than 200 EBP measures were identified in a recent umbrella review.^4^ In occupational therapy (OT) and physiotherapy (PT), there exist at least 30 self-report questionnaires which assess various EBP domains.^5–6^ Some questionnaires cover a single EBP domain such as self-efficacy (e.g., *Evidence-based Practice Confidence Scale*^7^) or attitudes (e.g., *Questions on EBP questionnaire*^8^) while others^9–22^ combine multiple domains in various groupings such as the *Evidence-Based Practice Profile Questionnaire* (EBP^2^).^23^

Given the various individual and organizational factors which, alone or in combination, may influence whether a clinician will integrate research evidence into CDM, it has been suggested to include all key EBP domains into a single index to gain a more comprehensive interpretation of EBP.^6,24^ Among the EBP measures recommended for use in rehabilitation derived from three systematic reviews,^11,12,31^ none comprehensively cover all domains associated with EBP, namely knowledge, attitudes, skills and behaviors^4^ and none measured the organizational factors that may influence EBP.^6^ This failure to combine domains for an overall index of EBP is one of the major shortcomings with existing EBP measures given that, in practice, all influencing factors may impact to varying degrees a clinician’s decision-making process. Since the publication of the three systematic reviews between 2010 and 2016,^5,6,25^ new methods for measuring multidimensional EBP in rehabilitation have been published and include items assessing the organizational-level factors known to influence EBP (e.g., the *Health Sciences Evidence Based Questionnaire*^9^ and the measure by Al Zoubi et al.^26^

An important challenge with many EBP questionnaires lies in the incorrect treatment of ordinal data. Most EBP questionnaires consist of ordinal scales whereby respondents select textual responses rather than numbers on a scale. Often, numbers are attributed to the ordinal response options and authors perform arithmetic operations on ordinal data (e.g., average across items).^27,28^ For instance, the EBP^2^ includes 74 items, covering five domains, all of which used a five-point Likert scale (e.g., not at all true, not really true, possibly true, quite likely true, very true).^23^ In one analysis,^29^ the ordinal data are treated as interval-level data as the authors present mean scores, standard deviations and effect sizes. Such an approach to analysis is flawed because the allocation of numbers is not indicative of any mathematically valid magnitude of difference between categories which can produce misleading conclusions.^28,30^

This work is part of a larger longitudinal study which aimed to measure and understand the evolution EBP competencies in OT and PT students from graduation and three years into practice.^26,31,32,33^ Given the limitations in the measurement of EBP competencies and the many existing measures of separate constructs, our research team conducted additional measurement work to develop a robust measurement approach for constructs around EBP including practice, individual (e.g. knowledge, attitudes, confidence, behaviors), and contextual factors (e.g. resources). Specifically, Al Zoubi and colleagues^26^ used Rasch analysis to determine the mathematical properties of six unidimensional scales of EBP. Items for each EBP domain were calibrated onto the same linear scale which allowed for a mathematically valid approach to summing scores. The measure developed by Al Zoubi et al^26^ has interval-level properties and resulted in six core EBP domains (knowledge, use of research evidence, self-efficacy, resources, attitudes, and activities related to EBP). The measure was them used in a four-year longitudinal study of OTs and PTs in Canada.^31,32,33^

Despite these recent developments in the measurement of EBP, separate values are generated for each domain, without the possibility of having one value covering the multidimensional nature of EBP. Considering that there is always a need for parsimony in measurement, a concise yet comprehensive measure able to generate an overall index of EBP in rehabilitation is needed.

### Objectives

The global aim of this study is to describe the prototype development of the *Propensity to Integrate Research Evidence into Clinical Decision-Making Index* (PIRE-CDMI). The specific study objectives are to (1) identify candidate items best reflecting the most salient EBP domains for OTs and PTs across Canada from existing measures; (2) estimate the extent to which the prototype PIRE-CDMI “behaves” coherently across characteristics of the sample; and (3) estimate the extent to which the prototype PIRE-CDMI provides information that is comparable to that from other EBP measures. As it will not have been subjected to qualitative revision or weight elicitation methods, this descriptive system will be referred to as the prototype index or the P-PIRE-CDMI. This study is designed to provide evidence to justify the need to develop a deployable measure.

## Methods

### Conceptual framework

Measuring a clinician’s *propensity* to integrate research evidence into CDM may be one approach to estimating and inferring EBP. We acknowledge that despite the original intentions of EBP, this approach to CDM is now viewed as acknowledging the contribution of more than just research evidence.^34^ Indeed, other components such as professional expertise, expert opinion, and patient preferences and values are recognized as contributing to the EBP process.^35–40^ To avoid a misnomer, this index is deliberately focused on the integration of research evidence which may include, but is not restricted to, peer-reviewed articles of original studies, systematic reviews and best practice guidelines).

In a conceptual model where *EBP* is the exposure and *improved patient health* is the outcome, propensity variables predict the exposure (i.e., EBP).^41^ In this study, *propensity* is described as a rehabilitation clinician’s conditional likelihood of integrating research evidence into CDM. The term propensity is defined by the Cambridge Academic Content Dictionary as “a tendency to behave in a particular way”. In applied statistics, propensity modeling is a set of techniques which attempts to estimate the likelihood of subjects performing a behavior by accounting for variables that affect such behavior. Using propensity as a proxy for behavior acknowledges the dynamic and uncertain nature of EBP and provides a practically relevant approach to measuring a complex construct.

There exists a well-articulated body of knowledge with respect to the influence of certain individual and organizational factors on the integration of research evidence into CDM and the measurement of those factors.^3,5–10,13–22,26,29,31,32,42^ This study will use the previously identified six core EBP domains in rehabilitation as the propensity variables (knowledge, use of research evidence, self-efficacy, resources, attitudes, and activities related to EBP).^26^ The underlying measurement model for the construct of propensity is a formative one (i.e., individual and organizational factors collectively produce the latent phenomenon).^26,43^ As such, the most appropriate approach in generating a total score for formative constructs is a weighted sum of dimensions, assuming that the dimensions are independent.

### Data source

This study consists of a secondary analysis of a dataset that sought to measure use of EBP each year over four years in newly graduated OTs and PTs.^26,31^ The detailed process for developing the multidimensional measure is described elsewhere.^26^

At baseline (T0), new graduates responded to this multidimensional 70-item online survey. Rasch analysis was conducted on their responses which allowed for item reduction by fixing or omitting certain items that did not fit the Rasch model. Rasch analysis can assess whether items span the entire construct, the extent to which items have response choices that are appropriately ordered and whether items perform differently between groups of people.^44–49^ The three subsequent surveys contained 55 items stemming from the six EBP domains - *Knowledge* (8 items, 0-29 scale), *Use of research evidence* (9 items, 5-9), *Self-efficacy* (8 items, 0-22), *Resources* (13 items. 0-39), *Attitudes* (10 items, 0-32), and *Activities related to EBP* (7 items, 1-5).

#### Sample characteristics

The sample used for analysis consisted of newly graduated OTs and PTs from 13 Canadian rehabilitation programs who were working at the time of survey completion and completed all items of the survey at baseline, T0 (n=127) and three years later, T3 (n=37). Table 1 summarizes the characteristics of the sample (mean age: 27; SD: 2.7-3.9, predominantly women).

**Table 1:**
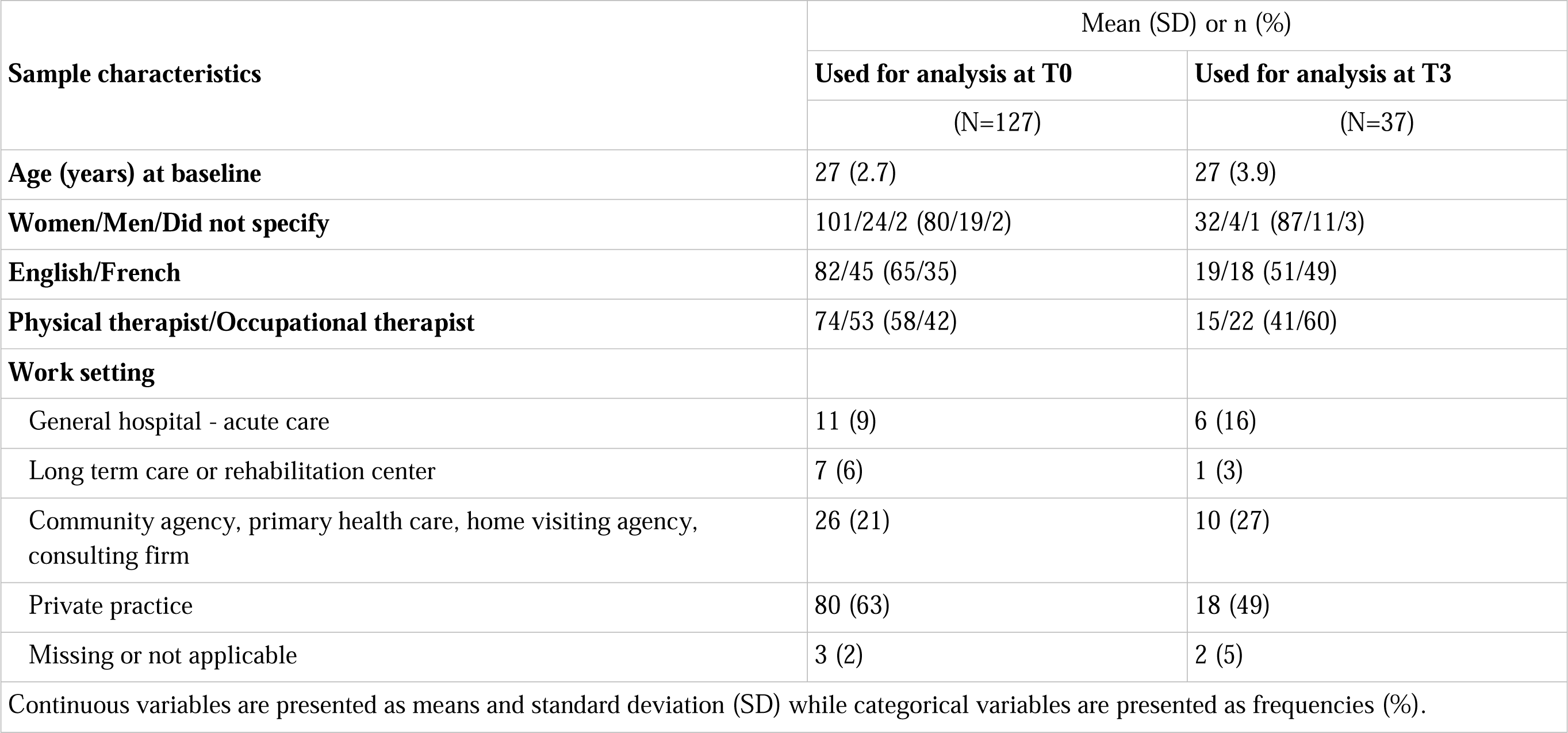
Descriptive demographics of the sample at baseline and three years later.

| Sample characteristics | Mean (SD) or n (%) |  |
| --- | --- | --- |
|  | Used for analysis at T0 | Used for analysis at T3 |
|  | (N=127) | (N=37) |
| Age (years) at baseline | 27 (2.7) | 27 (3.9) |
| Women/Men/Did not specify | 101/24/2 (80/19/2) | 32/4/1 (87/11/3) |
| English/French | 82/45 (65/35) | 19/18 (51/49) |
| Physical therapist/Occupational therapist | 74/53 (58/42) | 15/22 (41/60) |
| Work setting |  |  |
| General hospital - acute care | 11 (9) | 6 (16) |
| Long term care or rehabilitation center | 7 (6) | 1 (3) |
| Community agency, primary health care, home visiting agency, consulting firm | 26 (21) | 10 (27) |
| Private practice | 80 (63) | 18 (49) |
| Missing or not applicable | 3 (2) | 2 (5) |
| Continuous variables are presented as means and standard deviation (SD) while categorical variables are presented as frequencies (%). |  |  |

### Data analysis and procedure

#### A priori objectives for developing the prototype

We set out to develop a concise multidimensional measure which would reduce administration time and response burden for clinicians using a well-recognized method to develop preference-based measures. Our aim of selecting one item per domain is consistent with the standard way of reducing dimensions used in developing the Short-Form Six-Dimension or SF-6D and other preference-based measures for patients.^44–47,50–52^ The main advantage of such measures is their ability to generate an overall score that balances gains in one domain versus losses in another.^50–52^

#### Item selection

To build on the Rasch analysis threshold maps developed by Al Zoubi et al^26^ using the standard procedure for developing preference-based measures,^44,53^ two team members (JRD, NM) selected one item per reflective construct. Specifically, the best performing item per EBP domain had interval-like response options, spanned the continuum, and centered on logit zero. The remaining expert panel team members (AT, KS, AR) considered the theoretical relevance and representativeness, clarity, length, directionality, distribution of responses and potential biases of all candidate items.^54^ N.B. Selected items best representing the EBP domain are hereafter called *dimensions.*^46^

#### Development of a scoring algorithm

A scoring algorithm was developed to obtain a score for each participant and test the performance of the prototype index. As per the scaled score method,^53^ the logit placements on the Rasch continuum were used to develop preliminary weights that represented the interval spacing of each response option. To obtain a total score, the points associated with participants’ response for each of the five dimensions were summed. These weights are not to be confused with preference weights as participant preferences have not been elicited. Total scores were transformed to a 0-100 scale to ease interpretation and comparisons with the initial EBP measures which were also transformed onto 100. On the prototype index, zero represents neutral propensity to integrate research evidence into CDM and 100 represents the highest propensity.

Polychoric correlation coefficients were calculated to ensure that dimensions were not highly correlated with each other. We hypothesized that the correlation coefficients would be low to moderate (no greater than 0.6); otherwise, it may signal a need to eliminate one of the items to increase structural independence.

#### Analysis of the prototype across characteristics of the sample

Descriptive statistics were used to determine the distributional parameters for the P-PIRE-CDMI and the EBP measures. We assumed that a minimal important difference is half a SD of raw scores.^55^

Simple linear regression (SLR) was conducted to estimate the effect of single characteristics on the prototype score, at both time points. These characteristics were treated categorically: age (younger, middle, and older), gender (women, men, did not specify), profession (OT, PT) and setting (private practice, acute general hospital, long term care/rehabilitation center, and community/home visiting agency); age was also treated continuously and was centered for analysis.

Generalized estimating equation (GEE) analyses were conducted (exchangeable correlation structure) to determine if there were any important interactions which would signal a potential source of variance over time by characteristic (P-PIRE-CDMI score = [characteristic (age centered, age group, gender, profession, or setting) + time + characteristic*time]). GEE accounts for correlations in scores within the same individual at both timepoints (i.e., we fit time as the within-subject variable). The main effect of time was not interpreted in the model; we were interested in the potential interaction term.

#### Analysis of the prototype compared to EBP measures

Spearman Rho was used to correlate scores from the P-PIRE-CDMI and the five unidimensional EBP measures. It was hypothesized that measures would be moderately correlated (between 0.3 and 0.6) because the P-PIRE-CDMI is multidimensional, encompasses items included in the EBP measures and was completed by the same sample.

GEE analyses determined the extent to which average scores differed by measure. In this case, GEE was selected to control for the correlation from multiple measurements on the same individual (i.e., we fit measure as the within-subject variable).

To determine the extent to which the P-PIRE-CDMI provides information that is comparable to EBP measures across characteristics of the sample (age, gender, profession, setting or university), the data were analyzed parametrically and non-parametrically; this was conducted because of the ordinal nature of two EBP measures which violated the assumption of normality. First, GEE was conducted (Score = [measure + characteristic + measure*characteristic]). This analysis informed on whether there was a characteristic effect across all measures and whether the effect of measure depended on characteristic. All variables except for age were treated categorically. Second, an analysis of ranks using chi-square (χ^2^) was conducted to determine if the measures ranked characteristic groups differently.

To determine if the P-PIRE-CDMI behaves in a similar way to other measures over time, the standardized response mean (SRM) was calculated by dividing the mean difference in score from T3 to T0 by the SD of the mean difference.^56^ Cohen’s criteria were used to interpret the magnitude of the standardized response mean, where 0.2 is small, 0.5 is moderate, and 0.8 is large.^57^ Paired *t*-tests were also conducted to estimate the average difference at both timepoints for each measure. To account for the clustering of multiple measurements at the individual level at two time points, GEE was conducted to determine the presence of an interaction effect between measure and time (Score = [measure + time + measure*time]). Logistic regression was used to estimate whether age, gender, profession, setting, university, and language were associated with dropout rates from T0 to T3. SPSS Statistics Version 28 was used for all statistical analyses in this study.

## Results

### The prototype index

Five items were selected for inclusion in the P-PIRE-CDMI. The items, response options, threshold logit estimates, and score transformations are presented in Table 2. The polychoric correlations between included items are presented in Appendix 1 and ranged from 0.34 to 0.57.

**Table 2.** Candidate items, response options and score transformations.

| Dimension | Question | Selected item | Response options | Threshold logit scores | Threshold logit scores recalibrated | Recalibrated logit on 100 | Scoring on 10 |
| --- | --- | --- | --- | --- | --- | --- | --- |
| Use of research evidence | How often have you done each of the following activities in the past month? | Decide on an appropriate course of action based on integrating the research evidence, clinical judgment and patient or client preferences? | 0 (Never) | Formative model– no Rasch analysis conducted |  | 0 | 0 |
|  |  |  | 1 (One time or more) |  |  | 50 | 5 |
| Self-efficacy | Please indicate how confident you are in your current level of ability by choosing the corresponding number on the following rating scale. | Determine if the evidence from the research literature applies to your patient | 0 (0-25%) | NA | NA | 0 | 0 |
|  |  |  | 1 (25-50%) | -2.834 | 3.2 | 26.4 | 2.6 |
|  |  |  | 2 (50-75%) | -0.136 | 5.9 | 48.8 | 4.9 |
|  |  |  | 3 (75-100%) | 2.466 | 8.5 | 70.5 | 7 |
| Resources | Please indicate your level of agreement with the following statements. | My organization supports best practice | 0 (Strongly disagree /disagree) | NA | NA | 0 | 0 |
|  |  |  | 1 (Neutral) | -2.028 | 4.0 | 33.1 | 3.3 |
|  |  |  | 2 (Agree) | -0.274 | 5.7 | 47.7 | 4.8 |
|  |  |  | 3 (Strongly agree) | 1.845 | 7.8 | 65.3 | 6.5 |
| Attitudes | Please indicate your level of agreement with the following statements. | I am willing to use new and different types of clinical interventions (e.g., assessment, treatment) developed by researchers to help my patients/clients | 0 (Strongly disagree) | NA | bottom | 0 | 0 |
|  |  |  | 1 (Disagree) | -5.835 | 0.2 | 1.4 | 0 |
|  |  |  | 2 (Neutral) | -0.282 | 5.7 | 47.6 | 4.8 |
|  |  |  | 3 (Agree) | -0.114 | 5.9 | 49.0 | 4.8 |
|  |  |  | 4 (Strongly agree) | 1.74 | 7.7 | 64.5 | 6.5 |
| Activities related to EBP | In the past month, how often have you...? | Made time to read research | 0 (Never) | Formative model – no Rasch analysis conducted |  | 0 | 0 |
|  |  |  | 1 (Monthly or less) |  |  | 25 | 2.5 |
|  |  |  | 2 (Bi-weekly) |  |  | 50 | 5 |
|  |  |  | 3 (Weekly) |  |  | 75 | 7.5 |
|  |  |  | 4 (Daily) |  |  | 100 | 10 |

The expert panel omitted the *knowledge* domain because items in the initial measure only represented knowledge about statistical concepts which is not always necessary given the existence of evidence products (e.g., guidelines) and is not the only type of knowledge invoked when integrating research evidence into CDM (other types of knowledge include content knowledge of a best practice.^38,58^ Also, there is increasing recognition that knowledge is best assessed through cognitive testing not self-report.^59^ Indeed, a ceiling effect was found for the full *knowledge* measure wherein 84% of new graduates rated themselves very knowledgeable about statistical concepts.^26^

Appendix II presents the distribution of scores of EBP measures across characteristics of the sample at T0 (N=127), mean scores at T0 and T3 (n=37), average difference in means over time, standardized response mean and *t*-test results. No association was found between age, gender, profession, setting, university, and language with dropouts from T0 to T3.

### The prototype index across characteristics of the sample

Figures 1 to 4 illustrate the distribution of P-PIRE-CDMI scores across four characteristic groups at both timepoints. Appendix III presents the SLR estimates for the modeling of P-PIRE-CDMI scores as a function of characteristic, at both time points individually. Appendix IV reports the GEE estimates for the modeling of P-PIRE-CDMI scores as a function of characteristic, time, and the interaction.

**Figure 1.**
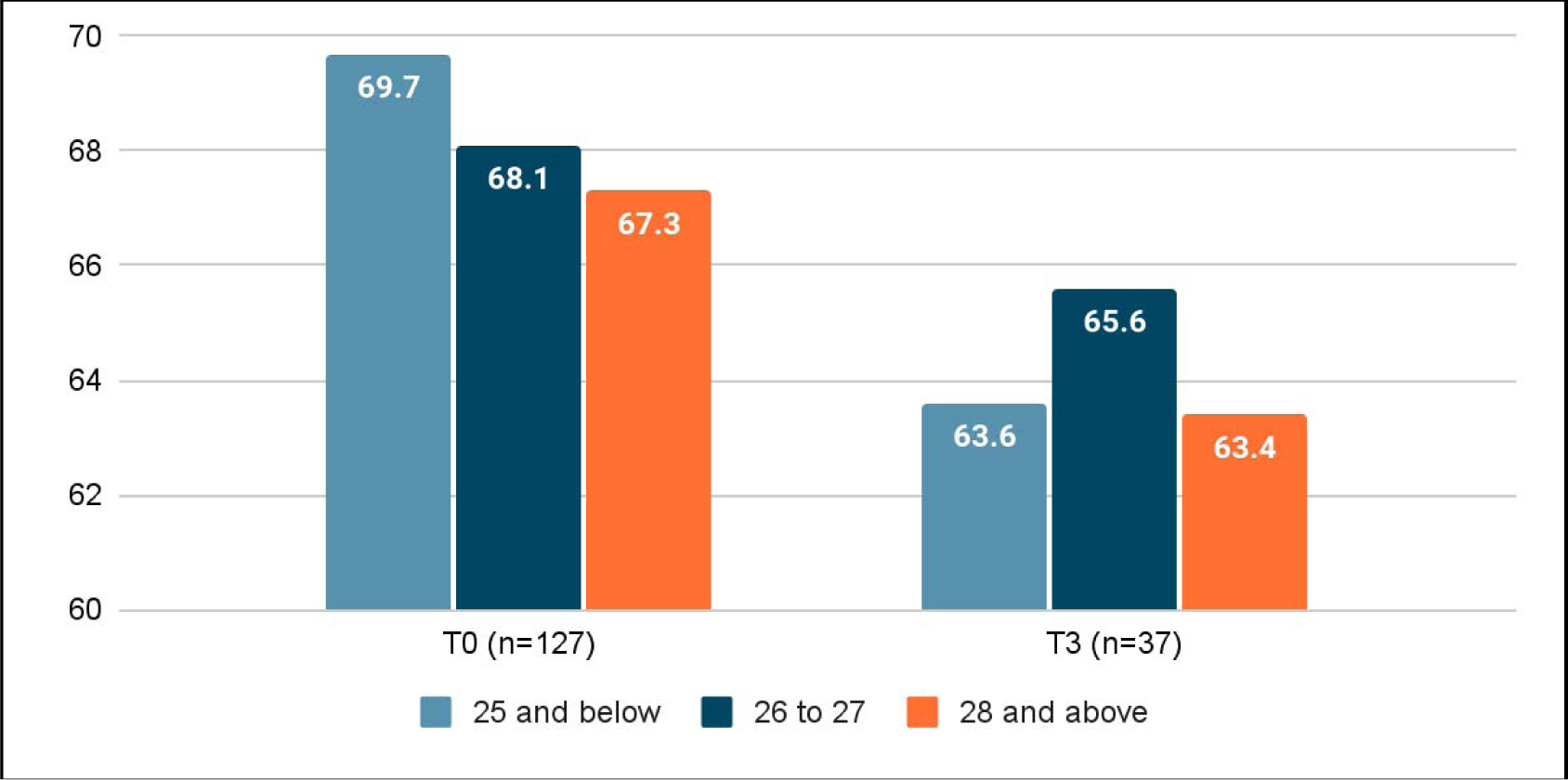
Distribution of prototype index scores across age groups at both timepoints

**Figure 2.**
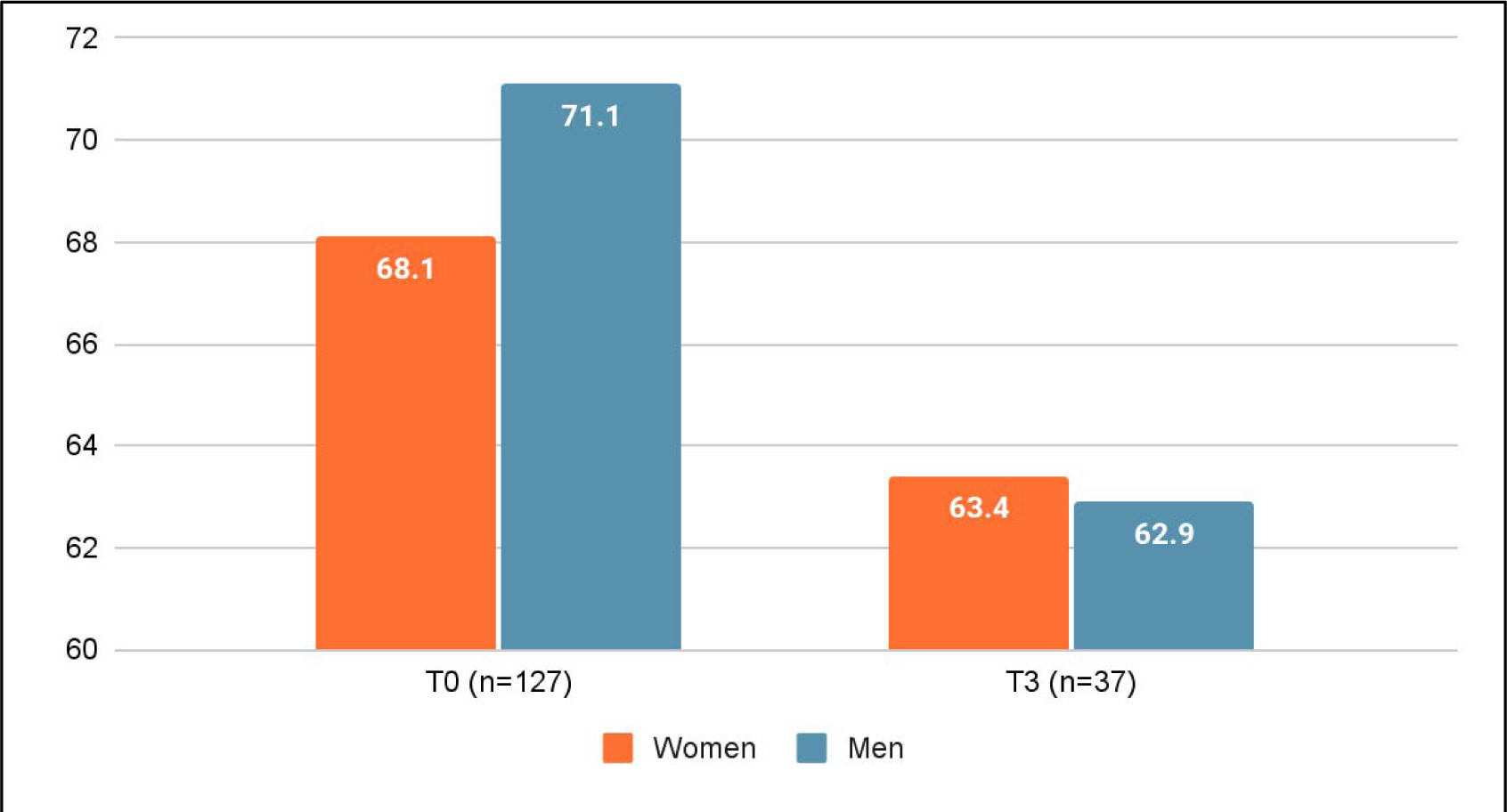
Distribution of prototype index scores across gender at both timepoints

**Figure 3.**
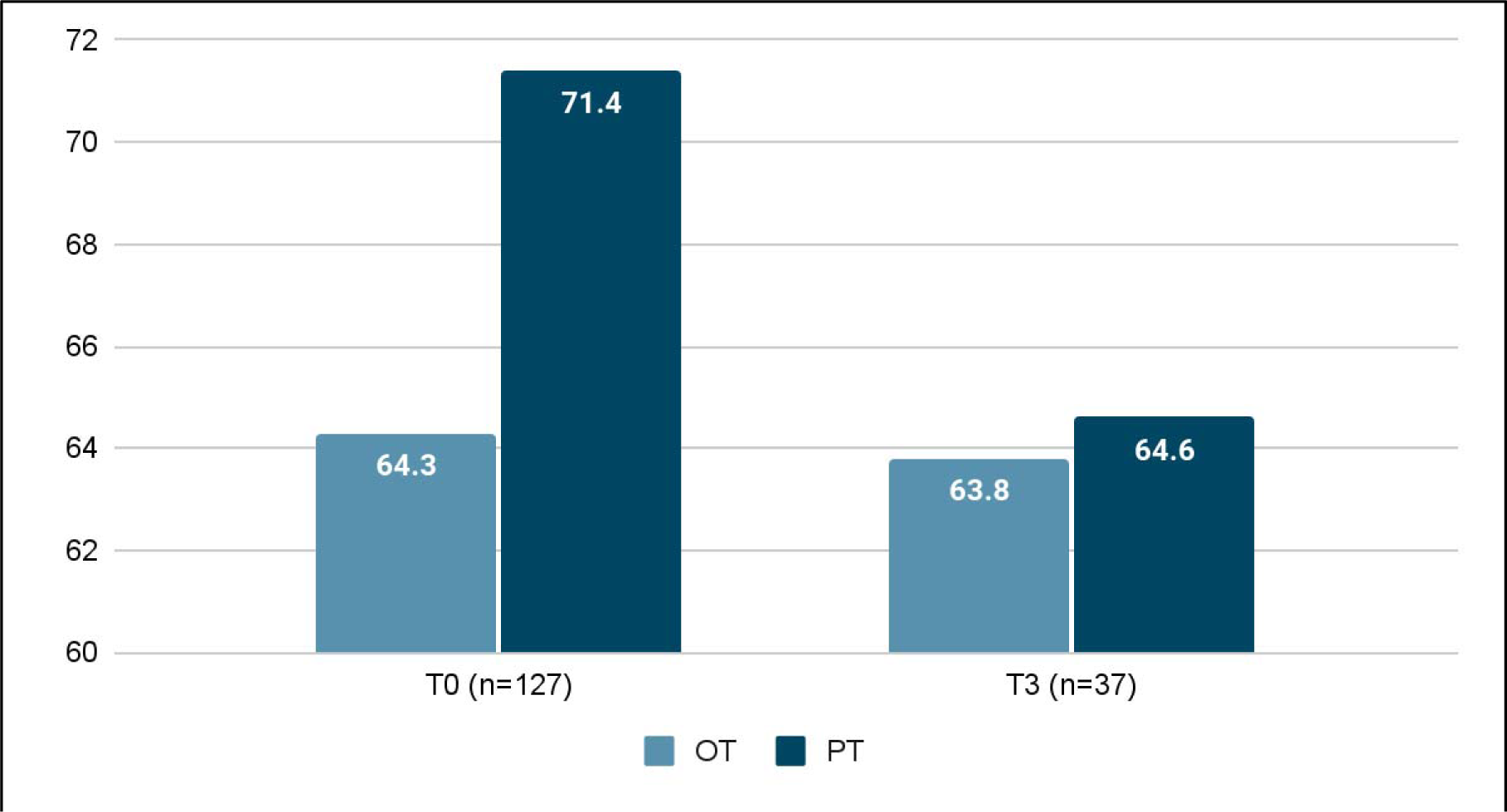
Distribution of prototype index scores across profession at both timepoints

**Figure 4.**
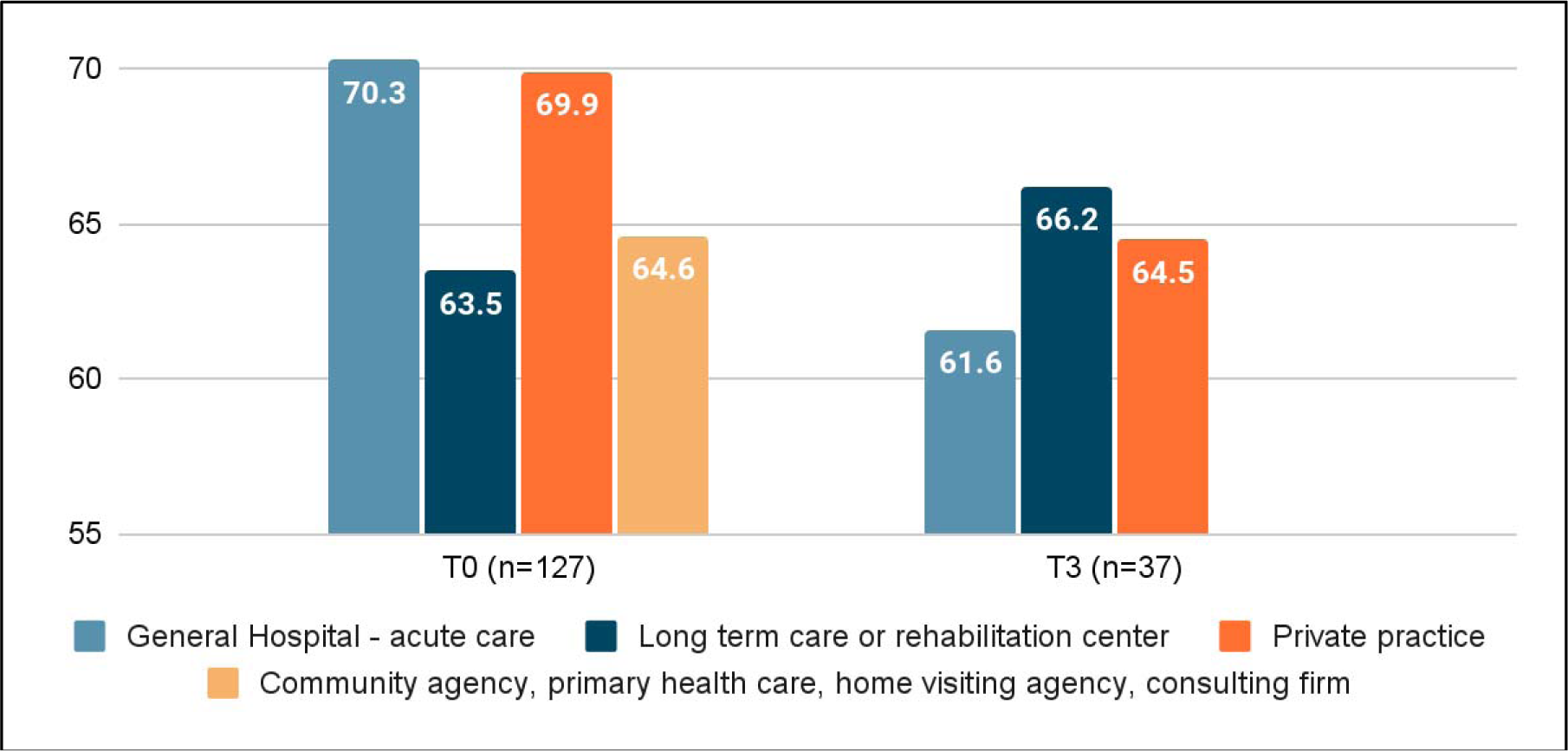
Distribution of prototype index scores across setting at both timepoints

There was no effect of gender or setting on P-PIRE-CDMI scores at both timepoints as suggested by SLR, and no important interaction of these individual characteristics with time in the GEE model. An important effect was found for profession at baseline whereby OTs scored, on average, 7.1 points lower on the P-PIRE-CDMI (95% CI: -11.5 to -2.6) compared to PTs. Three years later, there was no longer an effect of profession on P-PIRE-CDMI scores (β= -0.8, 95% CI: -9 to 7.4). GEE signaled an important interaction term of profession*time. SLR was suggestive of an age effect on P-PIRE-CDMI at baseline when treated continuously (β = -0.8, CI: -0.7 to 0.0). GEE was suggestive of an age effect when treated categorically (older age group: β = -11, CI: -19.6 to -2.4) and continuously (β = -1.6, CI: -2.2 to -0.9). After excluding three outliers from the sample of 127 participants at T0 and 37 participants at T3 (age at baseline: 36, 36 and 43), SLR no longer detected an effect of age at either time point, but GEE still found an effect of age when treated continuously (β= -1.7, CI: -3.4 to 0.0).

### The prototype index compared to other EBP measures

The P-PIRE-CDMI correlated moderately with the *self-efficacy* (r=0.36, [95% CI: 0.19 to 0.51]), *resources* (r=0.37, [95% CI: 0.21 to 0.52]), *use of research evidence* (r=0.46, [95% CI: 0.30 to 0.59]), *attitudes* (r=0.54, [95% CI: 0.40 to 0.65]) and *activities* (r=0.62, [95% CI: 0.49 to 0.72]) measures. Table 3 reports the GEE estimates illustrating the difference in average scores between measures at baseline.

**Table 3.** GEE estimates for scores as a function of EBP measure at baseline (N=127)

| | Estimate ($\beta$) | SE | 95% CI | | t-Statistic <sup>a</sup> ($\beta$ /SE) |
| --- | --- | --- | --- | --- | --- |
|  |  |  | Lower | Upper |  |
| **P-PIRE-CDMI** (referent) | 68.4 | 1.1 | 66.2 | 70.7 |  |
| **Activities** <sup>b</sup> | -55.7 | 1.2 | -57.9 | -53.4 | -48.1* |
| **Resources** | -3.6 | 1.4 | -6.2 | -0.9 | -2.6* |
| **Use of research evidence** | 11.9 | 1.4 | 9.1 | 14.7 | 8.4* |
| **Self-Efficacy** | -5.9 | 1.5 | -9.0 | -2.9 | -3.8* |
| **Attitudes** | 0.9 | 1.0 | -1.1 | 3.0 | 0.9 |
| GEE model: Score=measure |  |  |  |  |  |
| Dependent Variable: Score on each measure (out of 100) at baseline |  |  |  |  |  |
| GEE, generalized estimating equations; $\beta$, beta; SE, standard error; CI, confidence interval; P-PIRE-CDMI, prototype of the *Propensity to Integrate Research Evidence into Clinical Decision-Making Index* | | | | | |
| *There was an effect of the measure |  |  |  |  |  |
| <sup>a</sup> The t-statistic is a metric of effect size. |  |  |  |  |  |
| <sup>b</sup> Lower beta due to the transformation of the ordinal scale onto a 0-100 scale. |  |  |  |  |  |

Figures 5 to 8 portray the distribution of the P-PIRE-CDMI and five EBP measures by age group, gender, profession, and setting, respectively, at baseline.

**Figure 5.**
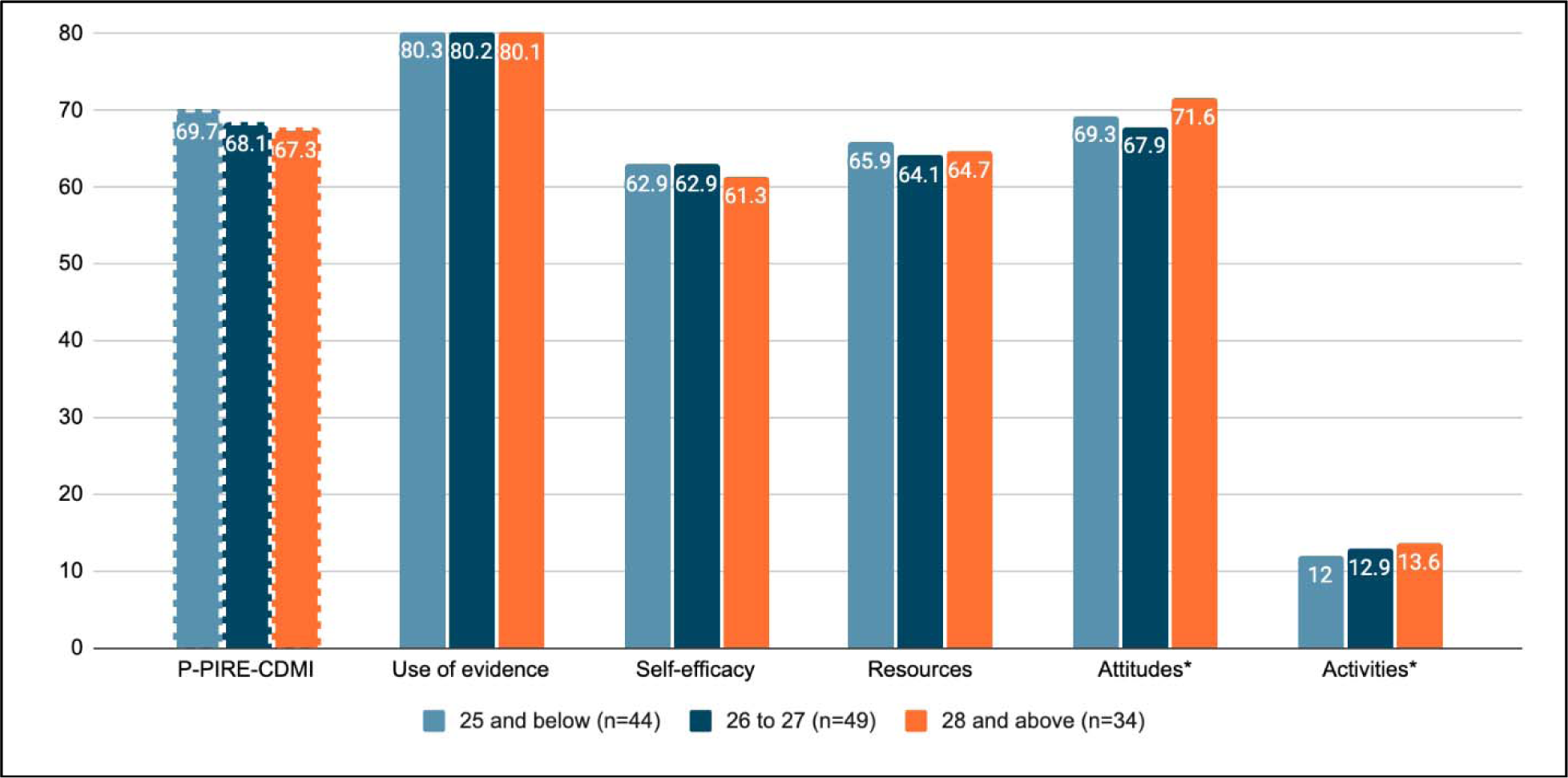
Distribution of scores from the prototype and five EBP measures by age group at baseline

**Figure 6.**
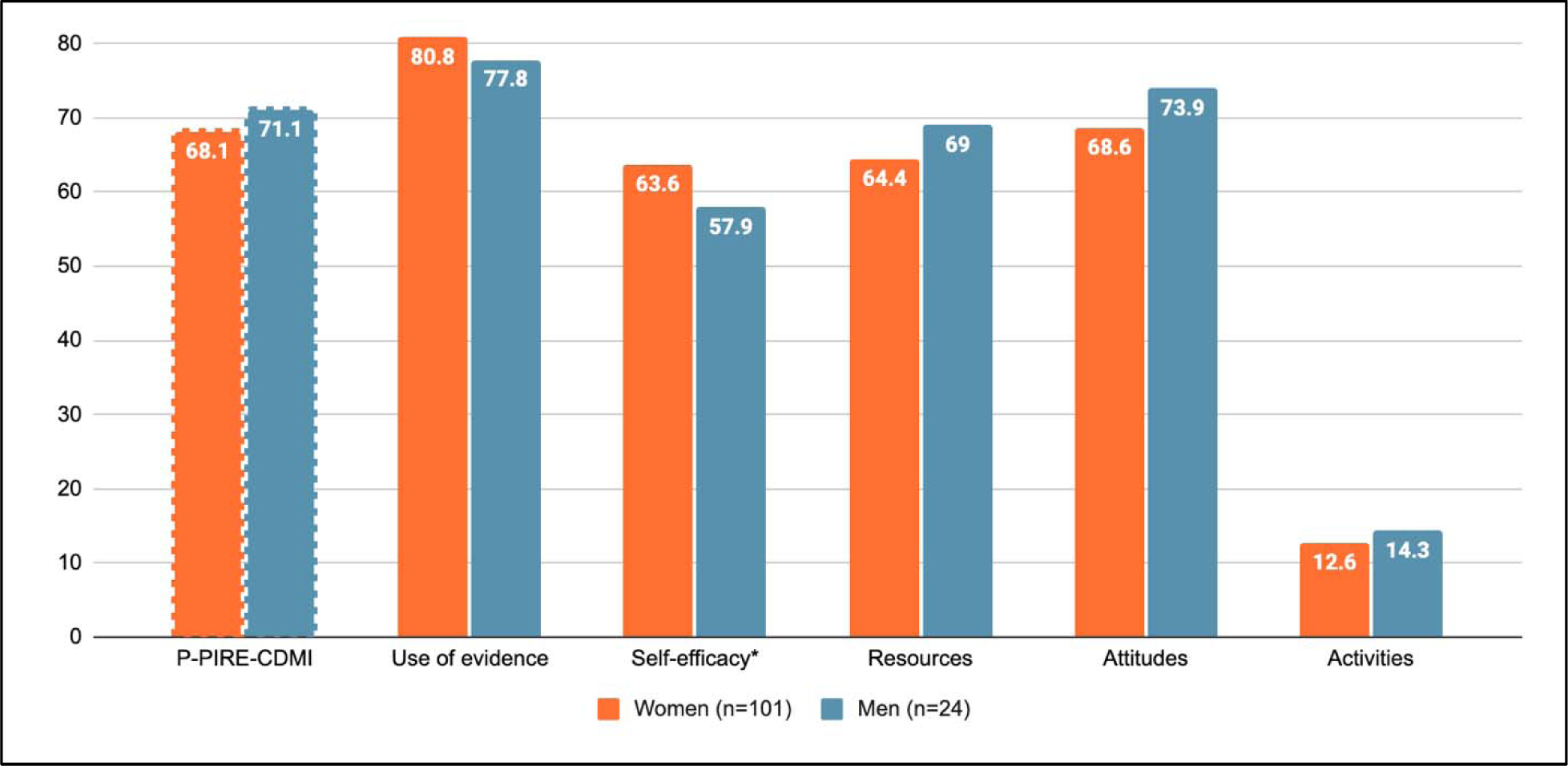
Distribution of scores from the prototype and five EBP measures by gender at baseline

**Figure 7.**
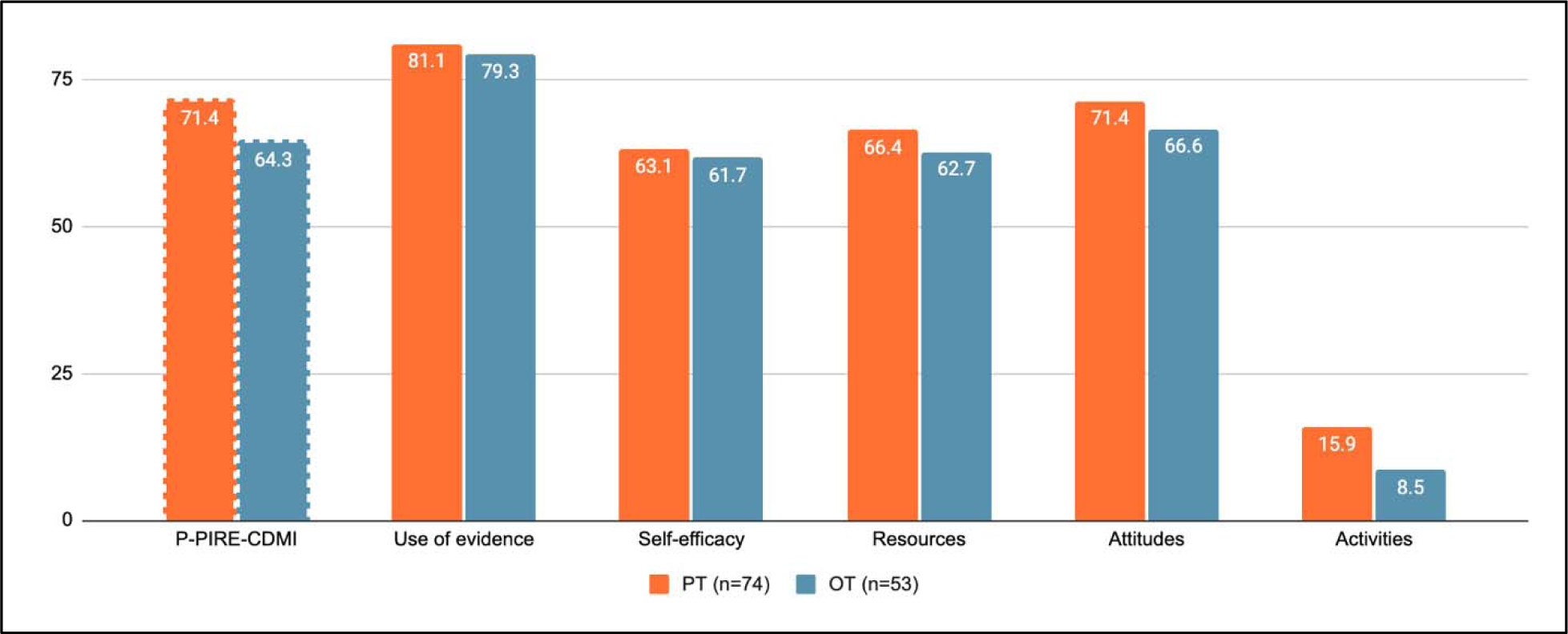
Distribution of scores from the prototype and five EBP measures by profession at baseline

**Figure 8.**
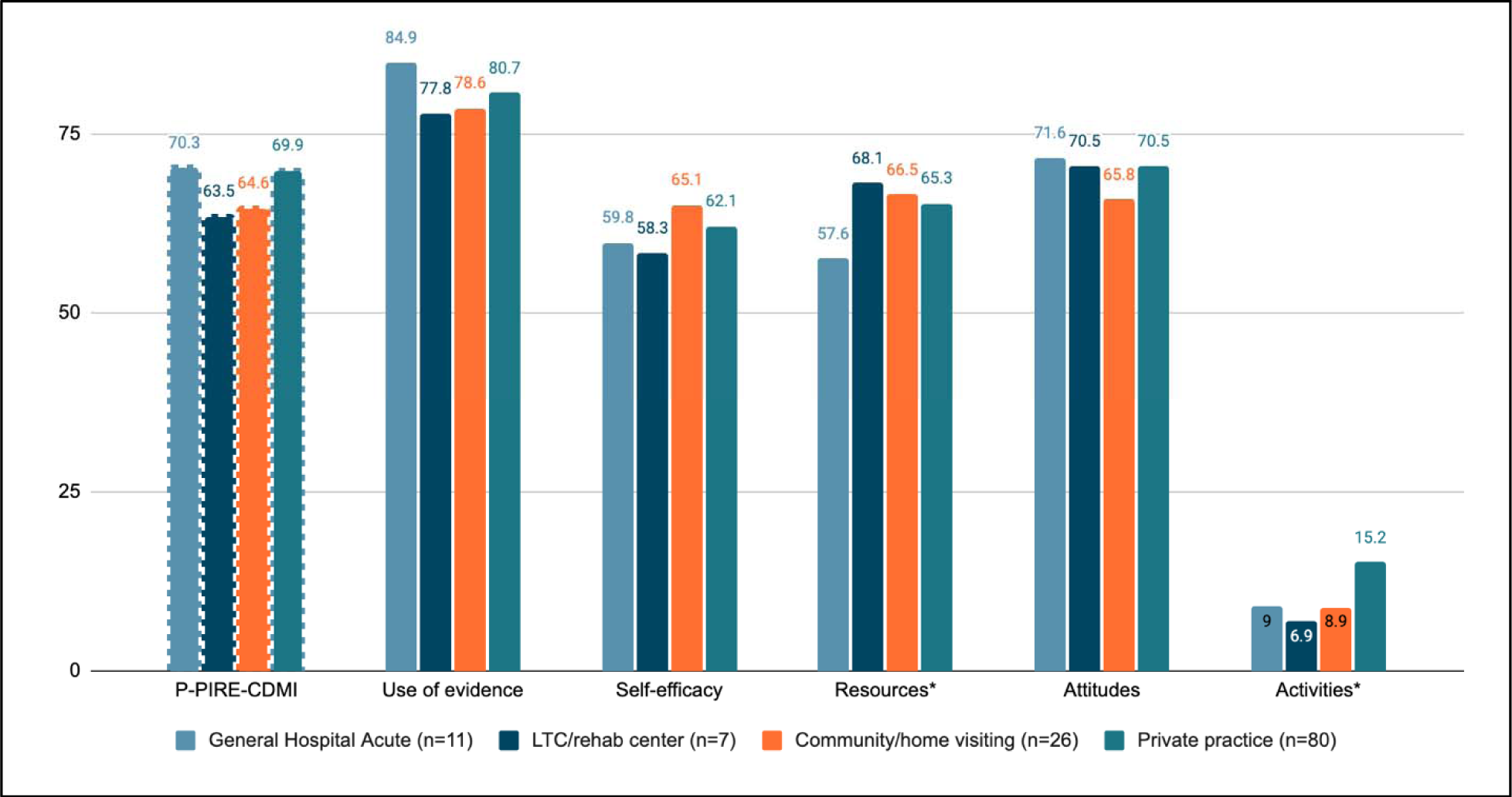
Distribution of scores from the prototype and five EBP measures by setting at baseline

There was an important effect of age (β: -0.85, 95% CI: -1.66 to -0.04) and profession across measures (β: -7.1, 95% CI: -11.5 to -2.6). Results suggests an important interaction effect of age with the *attitudes* (β: 1.1, 95% CI: 0.5 to 1.7) and *activities* (β: -0.87, 95% CI: 0.19 to 1.5) measures; gender with the *self-efficacy* (β: 8.7, 95% CI: 3.9 to 1.1) measure; setting with the *resources* (β: 8.1, 95% CI: 0.6 to 15.6 and β: 14.6, 95% CI: 5.8 to 23.3) and *activities* (β: 6.5, 95% CI: 0.1 to 13.0 and β: 5.6, 95% CI: -1.0 to 12.2) measures; and universities with the *self-efficacy* (β: 9.8, 95% CI: 0.4 to 19.2; β: -11.5, 95% CI: -21.3 to -1.8; β: -15.9, 95% CI: -27.8 to -3.9; and β: 11.5, 95% CI: 3.6 to 19.5) and *resources* (β: 13.3, 95% CI: 3.4 to 23.2) measures. No important difference in ranks of measures by age group (χ*^2^* (df:2, .05) =4.06, p=0.13), gender (χ^2^ (df:2, .05) =3.25, p=0.2) and profession (χ^2^ (df:2, .05) =0, p=1) was found. An important difference in ranks of measures by setting was found (χ^2^ (df:4, .05) =16.3, p=.003) and a very important heterogeneity in the ranking of universities by measure (χ^2^ (df:9, .05) =55.58, p=9.4E-9). Finally, our results did not find an important interaction effect of measure with time (i.e., there was no difference in the way measures performed from baseline to T3).

## Discussion

This study demonstrates how Rasch Measurement Theory and expert consensus can be leveraged to reduce 55 items to five items representing previously identified core EBP domains: (1) use of research evidence, (2) self-efficacy, (3) resources, (4) attitudes, and (5) activities related to EBP. Our specific study objectives were met by (1) identifying candidate items best reflecting key EBP domains for OTs and PTs across Canada from existing measures; (2) estimating the extent to which the prototype PIRE-CDMI “behaves” coherently across characteristics of the sample; and (3) estimating the extent to which the prototype PIRE-CDMI provides information that is comparable to that from other EBP measures. Our results provide promising evidence that a multidimensional index can offer a single score to rapidly identify a clinician’s propensity to integrate research evidence in their practice and highlight areas requiring further evaluation and support.

The low to moderate polychoric correlations between selected items support the posit that the items are related yet not redundant and that they measure separate domains of propensity to integrate research evidence into CDM.^43^ The expert panel omitted the knowledge dimension from the prototype index, in part, because they considered knowledge to be best assessed through cognitive testing and the items pertained to knowledge of statistical methods not knowledge about a specific evidence-based practice (assessment or treatment intervention). This decision is further justified given recent research demonstrating the absence of an association between knowledge scores and OTs’ and PTs’ self-reported use of research evidence.^31^ In addition, challenges with measuring the knowledge domain have been described in the literature and include low internal consistency,^60^ differential item functioning of knowledge items by ethnicity, gender and profession^61^ and overestimation of self-reported knowledge levels.^62^

Scores were normally distributed for the P-PIRE-CDMI without any floor or ceiling effects. The P-PIRE-CDMI behaved coherently across age, gender, and settings at both time points and there was no distinct source of variance on scores across these characteristics over time. There was an important difference in P-PIRE-CDMI scores between OTs and PTs at baseline whereby OTs scored 7 points lower on the index than PTs. An important profession effect was also found across all EBP measures at baseline. Though we have no way of knowing with certainty why this difference exists, it is possible that some complex elements of OT practice (e.g., environmental adaptations, occupational performance, or psychosocial approaches) may not be adequately addressed in the literature compared to components of PT practice.^63^ Our finding on the difference between OTs and PTs is consistent with a study on chronic pain by Arumugam et al.^64^ where OTs scored lowest in EBP behavior compared to other healthcare professionals including PTs. Our results confirm the importance of stratifying results related to EBP by rehabilitation profession given that these groups exhibit different trends.

Three years after baseline, there was no longer a difference between professions on P-PIRE-CDMI scores. PT scores dropped and OT scores remained stable. This decrease in scores may speak to a mechanism whereby clinical experience gained over time offsets the difference between professions as forms of knowledge other than that derived from empirical research become paramount.^32^ The decrease in EBP over time (as measured by the P-PIRE-CDMI and the five EBP measures) may be, in part, explained by variations in the definition of evidence.^65,66^ For new graduates, evidence may be closely related to knowledge gleaned from research articles and textbooks.^67^ With experience, the research-based knowledge once acquired through searching the literature becomes consolidated such that it may take the form of tacit or experiential knowledge and may no longer be distinguished as *evidence* or *research evidence*.^67–69^

As hypothesized, the P-PIRE-CDMI was moderately correlated with the five EBP measures. Our results suggest that the measure has an important effect on score distribution in terms of magnitude and direction. In other words, the value on the latent construct of EBP is inconsistent when using different measures. A combined interpretation of results from the five unidimensional EBP measures may be thwarted due to these diverging distributions. The P-PIRE-CDMI provides a single score that combines these five dimensions into one coherent indicator and behaves in a way that is consistent with the five EBP measures across time.

Furthermore, our findings reveal that when stratifying the sample by age, gender, setting and university, EBP measures provide very different perspectives. In contexts where researchers are seeking to determine EBP propensity as a function of university attended or type of clinical setting, we suggest considering the use of the PIRE-CDMI (future versions) as a first brief overall measurement which may then be complemented, when necessary, with in-depth measures of specific domains of interests.

Although we do not claim that the results of individual EBP measures are inaccurate, the combined interpretation of results from these measures may be challenging and ineffective due to the opposing distributions, variation in scores based on age, gender, setting and university and difference in rank ordering of settings and universities. The P-PIRE-CDMI presents the advantage of combining five dimensions into one succinct measure.

### Implications

Such an outcome measure could be used to identify EBP engagement levels in practice, create knowledge translation interventions and assess their effectiveness, or measure changes in EBP over time. Another potential use of the propensity score generated by such an index is for matching and stratification to account for group differences when estimating the effects of interventions in EBP studies. Clinicians could complete the index in a baseline questionnaire which could inform group allocation, sampling strategies or modelling through stratification by sub-groups based on propensity scores.

The prototype development described in this paper provides promising evidence that a practical five-item index offering a global interpretation of propensity for EBP could be deployed (pending further development on qualitative revision and the weighting of items) as an outcome measure in applied research. The continued development of this index responds to a call for multidimensional and pragmatic measures to support EBP research in rehabilitation.^24,70^ Pragmatic measures have been characterized as being low in burden for respondents and administrators, broadly applicable, actionable, and unlikely to cause harm.^70^ Another possible expansion of the index is as a self-reflection or professional development tool in clinical practice.

### Limitations

This study was designed to produce a proof-of-concept measure and to provide preliminary evidence justifying continuation of the PIRE-CDMI development process. Accordingly, we invite readers to acknowledge that more research is required to refine these P-PIRE-CDMI scores. The estimates of P-PIRE-CDMI scores are based on an additive formula of the logit placements which may be mathematically valid but may not be theoretically valid; to do so would require eliciting dimension weights based on the perspectives of end-users, a step which is planned in the development process. Third, the beta estimates using the GEE methods (with time as a factor) should be interpreted with the consideration that we only used two time points. We did not aim to estimate the extent to which real change occurred. A group-based trajectory modeling of EBP constructs using the same dataset is published elsewhere.^32^

We acknowledge that the EBP scores presented in this paper may be overestimates for three reasons. First, clinicians may provide responses in a more socially acceptable way because EBP is a professional expectation, and they may not be inclined to disagree with its value (i.e., social desirability bias). Second, there is a likelihood that the convenience sample used in this paper is more positively inclined towards EBP compared to non-respondents. Thirdly, the sample used in this study is composed of new graduates who have been found to demonstrate higher knowledge,^14^ attitudes,^14,16,71,72^ skills^14,72,73^ and behaviors^74^ compared to experienced clinicians.

## Conclusion

In this study, we identified one best performing item for each of the five core EBP domains using previously conducted Rasch analysis. These five items constitute the prototype *Propensity to Integrate Research Evidence into Clinical Decision-Making Index* (PIRE-CDMI) which combines five dimensions onto a single scale with a mathematically valid scoring system. Testing of the P-PIRE-CDMI demonstrates that it behaves consistently across age group, gender, and setting. Compared to other unidimensional EBP measures, the P-PIRE-CDMI provides comparable information in a succinct way. This study highlights the benefits of a brief, multidimensional index to assess an OT or PT’s propensity to integrate research evidence into CDM.

## Data Availability

The datasets generated and/or analysed during the current study are available from the corresponding author on reasonable request.

## Acknowledgments

This study was funded by the Canadian Institutes of Health Research (grant number 148544). The funding body did not influence the study results and was not involved in the study in any way.

## List of abbreviations

ß: Beta
CDM: Clinical decision-making
χ^2^: Chi-square
CI: Confidence interval
EBP: Evidence-based practice
GEE: Generalized estimating equation
OT: Occupational therapy
OTs: Occupational therapists
P-PIRE-CDMI: Prototype Propensity to Integrate Research Evidence into Clinical Decision-Making Index
PIRE-CDMI: Propensity to Integrate Research Evidence into Clinical Decision-Making Index
PT: Physiotherapy
PTs: Physical therapists
SLR: Simple linear regression
SRM: Standardized response mean

