## Appendices I-IV for "Identifying candidate items for a prototype index on propensity to integrate research evidence into clinical decision-making in rehabilitation"

### Appendix I: Prototype PIRE-CDMI inter-item polychoric correlation matrix

|  | Use of research evidence | Self-efficacy | Resources | Attitudes |
| --- | --- | --- | --- | --- |
| Self-efficacy | 0.17 |  |  |  |
| Resources | 0.34 | 0.39 |  |  |
| Attitudes | 0.26 | 0.35 | 0.57 |  |
| Activities | 0.12 | 0.34 | 0.30 | 0.47 |

### Appendix II. Distribution of scores of EBP measures across characteristics of the sample at T0 (N=127) and at T3 (n=37)

|  | **Measures of EBP at T0** | | | | | | | | | | | |
| --- | --- | --- | --- | --- | --- | --- | --- | --- | --- | --- | --- | --- |
| **Characteristics** | **P-PIRE-CDMI** | | **Use of research evidence** | | **Self-efficacy** | | **Resources** | | **Attitudes** | | **Activities** | |
|  | 5 items | | 9 items | | 8 items | | 13 items | | 10 items | | 7 items | |
|  | Mean | SD | Mean | SD | Mean | SD | Mean | SD | Mean | SD | Mean | SD |
| **AGE** | | | | | | | | | | | | |
| 25 and below (n=44) | 69.7 | 13.8 | 80.6 | 17.1 | 62.9 | 17.7 | 65.9 | 13.6 | 69.3 | 12.3 | 12 | 11.8 |
| 26 to 27 (n=49) | 68.1 | 12.8 | 80.2 | 16.4 | 62.9 | 16.9 | 64.1 | 13.7 | 67.9 | 12.2 | 12.9 | 13.1 |
| 28 and above (n=34) | 67.3 | 11.8 | 80.1 | 17.7 | 61.3 | 18.2 | 64.7 | 16.9 | 71.6 | 10.2 | 13.6 | 15.9 |
| **GENDER** ^a^ | | | | | | | | | | | | |
| Men (n=24) | 71.1 | 13.8 | 77.8 | 16.7 | 57.9 | 13.8 | 69 | 14.8 | 73.9 | 14 | 14.3 | 14.6 |
| Women (n=101) | 68.1 | 12.5 | 80.8 | 17 | 63.6 | 18.2 | 64.4 | 13.8 | 68.6 | 10.7 | 12.6 | 13.3 |
| **PROFESSION** | | | | | | | | | | | | |
| Physical therapist (n=74) | 71.4 | 11.9 | 81.1 | 16.8 | 63.1 | 16.9 | 66.4 | 13.4 | 71.4 | 11 | 15 .9 | 15.4 |
| Occupational therapist (n=53) | 64.3 | 13 | 79.3 | 17 | 61.7 | 18.3 | 62.7 | 15.8 | 66.6 | 12.2 | 8.5 | 8.5 |
| **SETTING** ^b^ | | | | | | | | | | | | |
| General Hospital - acute care (n=11) | 70.3 | 11.3 | 84.9 | 13.4 | 59.8 | 21.5 | 57.6 | 13.5 | 71.6 | 8.6 | 9 | 6.7 |
| Long term care or rehabilitation center (n=7) | 63.5 | 18.9 | 77.8 | 19.3 | 58.3 | 27.2 | 68.1 | 18.4 | 70.5 | 16.2 | 6.9 | 5.9 |
| Community agency, primary health care, home visiting agency, consulting firm (n=26) | 64.6 | 11.1 | 78.6 | 18.6 | 65.1 | 19.7 | 66.5 | 14.1 | 65.8 | 12.6 | 8.9 | 13.4 |
| Private practice (n=80) | 69.9 | 13 | 80.7 | 16.6 | 62.1 | 15.1 | 65.3 | 14.6 | 70.5 | 11.2 | 15.2 | 14.3 |
| **UNIVERSITY** | | | | | | | | | | | | |
| B (n=10) | 72.1 | 12 | 74.4 | 17.4 | 61.8 | 16.6 | 60.8 | 11.4 | 70.6 | 9 | 14.3 | 13 |
| C (n=18) | 70.2 | 13.6 | 87 | 18.4 | 75.2 | 19.5 | 60.5 | 11.2 | 68.1 | 12.2 | 17.6 | 16.4 |
| D (n=13) | 74.1 | 10.9 | 86.3 | 13.7 | 61.5 | 17.3 | 69.4 | 11.5 | 72.6 | 11.1 | 16 | 15.1 |
| E (n=9) | 72 | 13.3 | 85.2 | 13.6 | 49.5 | 11.5 | 65.2 | 15.8 | 70.5 | 12.5 | 13.7 | 15 |
| F (n=20) | 68 | 10.9 | 76.7 | 18.4 | 66.6 | 15.3 | 61.5 | 14.7 | 69.2 | 14.1 | 15.1 | 16.3 |
| G (n=7) | 73.3 | 7.2 | 84.1 | 18 | 55.2 | 16.7 | 67 | 13.7 | 77.2 | 9.7 | 18.9 | 19.2 |
| H (n=14) | 68.3 | 13.5 | 81 | 16.6 | 56.5 | 12.4 | 71.3 | 11.8 | 71 | 7 | 9 | 6 |
| I (n=5) | 68.4 | 12.3 | 86.7 | 14.5 | 55.5 | 13.4 | 71.8 | 11.8 | 70.6 | 13 | 13.4 | 12 |
| J (n=22) | 60.2 | 14.7 | 71.2 | 15.6 | 63.4 | 20.1 | 66.3 | 20.6 | 64.1 | 12.2 | 4.6 | 3.2 |
| Group of 6 universities ^c^ (n=9) | 66.7 | 11.5 | 82.7 | 13.7 | 60.1 | 13.2 | 59.5 | 11.8 | 69.1 | 12.7 | 11.2 | 10.5 |
| **TIME** | | | | | | | | | | | | |
| T0 (n=127) | 68.4 | 12.9 | 80.3 | 16.9 | 62.5 | 17.4 | 64.9 | 14.5 | 69.4 | 11.7 | 12.8 | 13.4 |
| T0 with follow-up at T3 (n=37) | 67.8 | 12.8 | 77.8 | 17.4 | 67.5 | 20.1 | 63.3 | 16.3 | 70.2 | 9 | 12 | 12 |
| T3 (n=37) | 64.2 | 11.9 | 71.8 | 15.4 | 61.8 | 15.1 | 63.8 | 13.7 | 63.2 | 9.7 | 7.9 | 7 |
| Difference T3-T0 (n=37) | -3.7 | 14.3 | -6 | 19.9 | -5.7 | 14.5 | 0.4 | 17.6 | -7 | 12.3 | -4.1 | 10.4 |
| SRM ^d^ | -0.26 |  | -0.3 |  | -0.39 |  | 0.02 |  | -0.6 |  | -0.39 |  |
| Paired t-test (95% CI, df: 36) | -1.57 |  | -1.84 |  | -2.38 |  | 0.14 |  | -3.47 |  | -2.4 |  |
| **RANGE** ^e^ (min, max, range) | | | | | | | | | | | | |
| T0 (n=127) | 42 | 95.1 | 55.6 | 100 | 13.6 | 99.1 | 20.5 | 94.9 | 37.5 | 96.9 | 0 | 75 |
| T0 with follow-up at T3 (n=37) | 42.6 | 95.1 | 55.6 | 100 | 22.7 | 99.1 | 20.5 | 94.9 | 53.1 | 96.9 | 0.7 | 50 |
| T3 (n=37) | 41.4 | 92.9 | 55.6 | 100 | 31.8 | 95.5 | 35.9 | 94.9 | 43.8 | 87.5 | 0.7 | 25 |
| ^a^ No reporting for the *did not specify* group because n=2 (less than 5)  ^b^ No reporting for the *Missing or N‎/A* group because n=3 (less than 5)  ^c^ Six universities were grouped together (A, K, L, M, N, O) because sample sizes were less than 5 per university  ^d^ SRM is a metric of effect size  ^e^ The theoretical range for each measure is 0-100.  EBP, evidence-based practice; P-PIRE-CDMI**,** prototype of the *Propensity to Integrate Research Evidence into Clinical Decision-Making Index;* SD, standard deviation; SRM, standardized response mean calculated as (T3- T0)/SD of the group's score differences; CI, confidence interval; df, degrees of freedom | | | | | | | | | | | | |

### Appendix III. Regression estimates of the effect of characteristic on P-PIRE-CDMI scores at T0 and T3

| P-PIRE-CDMI score = characteristic | **T0** | | | | **T3** | | | |
| --- | --- | --- | --- | --- | --- | --- | --- | --- |
|  | **Unstandardized Coefficients** | | **95% CI for ß** | | **Unstandardized Coefficients** | | **95% CI for ß** | |
|  | Estimate (ß) | SE | Lower Bound | Upper Bound | Estimate (ß) | SE | Lower Bound | Upper Bound |
| **AGE-** *CATEGORICAL* |  |  |  |  |  |  |  |  |
| *25 and below (n_T0_=44; n_T3_=16) | 69.7 | 1.9 | 65.9 | 73.6 | 63.6 | 3.1 | 57.4 | 69.8 |
| 26,27 (n_T0_=49; n_T3_=11) | -1.6 | 2.7 | -6.9 | 3.7 | 2.0 | 4.8 | -7.7 | 11.8 |
| 28 above (n_T0_=34; n_T3_=10) | -2.4 | 2.9 | -8.3 | 3.4 | -0.2 | 4.9 | -10.2 | 9.8 |
| ***3 outliers excluded*** |  |  |  |  |  |  |  |  |
| *25 and below (n_T0_=44; n_T3_=16) | 69.7 | 1.9 | 65.9 | 73.5 | n=16 | 63.6 | <.001 | 58.0 |
| 26,27 (n_T0_=49; n_T3_=11) | -1.6 | 2.6 | -6.8 | 3.6 | n=11 | 2.0 | 0.639 | -6.7 |
| 28 above (n_T0_=31; n_T3_=7) | -0.7 | 3.0 | -6.6 | 5.2 | n=7 | -0.3 | 0.945 | -10.5 |
| **AGE -** *CONTINOUS* |  |  |  |  |  |  |  |  |
| Constant | 91.2 | 11.4 | 68.6 | 113.9 |  | 75.5 | <.001 | 47.0 |
| Age at Time 0 (n_T0_=127; n_T3_=37) | -0.8 | 0.4 | -1.7 | 0.0 | n=37 | -0.4 | 0.421 | -1.5 |
| ***3 outliers excluded*** |  |  |  |  |  |  |  |  |
| Constant | 114.4 | 30.7 | 51.9 | 176.8 |  | 79.9 | 0.008 | 22.2 |
| Age at Time 0 (n_T0_=124; n_T3_=34) | -1.7 | 1.2 | -4.1 | 0.7 | n=34 | -0.6 | 0.582 | -2.8 |
| **GENDER** |  |  |  |  |  |  |  |  |
| *Men (n_T0_=24; n_T3_=4) | 71.1 | 2.6 | 66.0 | 76.3 | 62.9 | 5.6 | 51.5 | 74.3 |
| Women (n_T0_=101; n_T3_=32) | -3.0 | 2.9 | -8.7 | 2.7 | 0.5 | 5.9 | -11.6 | 12.6 |
| **PROFESSION** |  |  |  |  |  |  |  |  |
| *Physical therapist (n_T0_=74; n_T3_=15) | 71.4 | 1.4 | 68.5 | 74.2 | 64.6 | 3.1 | 58.3 | 71.0 |
| Occupational therapist (n_T0_=53; n_T3_=22) | -7.1 | 2.2 | -11.5 | -2.6 | -0.8 | 4.0 | -9.0 | 7.4 |
| **SETTING** |  |  |  |  |  |  |  |  |
| *Private practice (n_T0_=80; n_T3_=18) | 69.9 | 1.4 | 67.1 | 72.8 | 64.5 | 2.9 | 58.6 | 70.4 |
| General Hospital Acute (n_T0_=11; n_T3_=6) | 0.4 | 4.1 | -7.8 | 8.5 | -2.9 | 5.8 | -14.7 | 8.9 |
| Long term care/rehabilitation center (n_T0_=7; n_T3_=1) | -6.4 | 5.1 | -16.4 | 3.6 |  |  |  |  |
| Community agency, primary health care, home visiting agency, consulting firm (n_T0_=26; n_T3_=10) | -5.3 | 2.9 | -11.0 | 0.4 | 1.7 | 4.8 | -8.2 | 11.6 |
| *referent category  P-PIRE-CDMI**,** prototype of the *Propensity to Integrate Research Evidence into Clinical Decision-Making Index;* *ß,* beta; SE, standard error; CI, confidence interval | | | | | | | | |

### Appendix IV: GEE estimates of the effects of characteristic and time on P-PIRE-CDMI scores

1) Age categorical with and without outliers (group 1 = 25 and below; group 2 = 26 and 27; group 3 = 28 and above)

**P-PIRE-CDMI =age (categorical) + time + interaction (Age group*Time)**

| Parameter Estimates (with outliers, n=37) | | | | | | | |
| --- | --- | --- | --- | --- | --- | --- | --- |
| Parameter | Estimate (ß) | Standard error | 95% Wald Confidence Interval | | Wald Chi-Square | df | Sig. |
|  |  |  | Lower | Upper |  |  |  |
| (Intercept) | 71.3 | 3.2 | 64.9 | 77.7 | 482.3 | 1 | 0 |
| [time=2] | -7.7 | 2.9 | -13.3 | -2.1 | 7.1 | 1 | 0.008 |
| [time=1] | 0a | . | . | . | . | . | . |
| [age_group=3] | -11.0 | 4.4 | -19.6 | -2.4 | 6.2 | 1 | 0.012 |
| [age_group=2] | -1.7 | 4.8 | -11.2 | 7.7 | 0.1 | 1 | 0.721 |
| [age_group=1] | 0a | . | . | . | . | . | . |
| [time=2] * [age_group=3] | 10.8 | 6.1 | -1.1 | 22.7 | 3.2 | 1 | 0.076 |
| [time=2] * [age_group=2] | 3.7 | 4.7 | -5.4 | 12.9 | 0.6 | 1 | 0.421 |
| [time=2] * [age_group=1] | 0a | . | . | . | . | . | . |
| [time=1] * [age_group=3] | 0a | . | . | . | . | . | . |
| [time=1] * [age_group=2] | 0a | . | . | . | . | . | . |
| [time=1] * [age_group=1] | 0a | . | . | . | . | . | . |
| Dependent Variable: P-PIRE-CDMI T0 100 | | | | | | | |

| Parameter Estimates (without outliers, n=34) | | | | | | | |
| --- | --- | --- | --- | --- | --- | --- | --- |
| Parameter | Estimate (ß) | Standard error | 95% Wald Confidence Interval | | Wald Chi-Square | df | Sig. |
|  |  |  | Lower | Upper |  |  |  |
| (Intercept) | 71.3 | 3.2 | 64.9 | 77.7 | 482.3 | 1 | 0 |
| [time=2] | -7.7 | 2.9 | -13.3 | -2.1 | 7.1 | 1 | 0.008 |
| [time=1] | 0a | . | . | . | . | . | . |
| [age_group=3] | -6.3 | 3.9 | -13.9 | 1.3 | 2.7 | 1 | 0.102 |
| [age_group=2] | -1.7 | 4.8 | -11.2 | 7.7 | 0.1 | 1 | 0.721 |
| [age_group=1] | 0a | . | . | . | . | . | . |
| [time=2] * [age_group=3] | 6.0 | 4.0 | -1.8 | 13.8 | 2.2 | 1 | 0.134 |
| [time=2] * [age_group=2] | 3.7 | 4.7 | -5.4 | 12.9 | 0.6 | 1 | 0.421 |
| [time=2] * [age_group=1] | 0a | . | . | . | . | . | . |
| [time=1] * [age_group=3] | 0a | . | . | . | . | . | . |
| [time=1] * [age_group=2] | 0a | . | . | . | . | . | . |
| [time=1] * [age_group=1] | 0a | . | . | . | . | . | . |
| Dependent Variable: P-PIRE-CDMI T0 100 | | | | | | | |

2) Age continuous with and without outliers

**P-PIRE-CDMI =age (continuous) + time + interaction (Age *Time)**

| Parameter Estimates (with outliers, n=37) | | | | | | | |
| --- | --- | --- | --- | --- | --- | --- | --- |
| Parameter | Estimate (ß) | Standard error | 95% Wald Confidence Interval | | Wald Chi-Square | df | Sig. |
|  |  |  | Lower | Upper |  |  |  |
| (Intercept) | 111.0 | 9.9 | 91.6 | 130.3 | 126.5 | 1 | 0 |
| [time=2] | -35.5 | 24.7 | -83.8 | 12.8 | 2.1 | 1 | 0.15 |
| [time=1] | 0a | . | . | . | . | . | . |
| Age at T0 | -1.6 | 0.3 | -2.2 | -0.9 | 23.2 | 1 | <.001 |
| [time=2] * Age at Time 0 | 1.2 | 0.9 | -0.7 | 3.0 | 1.5 | 1 | 0.213 |
| [time=1] * Age at Time 0 | 0a | . | . | . | . | . | . |
| Dependent Variable: P-PIRE-CDMI T0 100 | | | | | | | |

| Parameter Estimates (without outliers, n=34) | | | | | | | |
| --- | --- | --- | --- | --- | --- | --- | --- |
| Parameter | Estimate (ß) | Standard error | 95% Wald Confidence Interval | | Wald Chi-Square | df | Sig. |
|  |  |  | Lower | Upper |  |  |  |
| (Intercept) | 114.4 | 23.6 | 68.1 | 160.6 | 23.5 | 1 | 0 |
| [time=2] | -34.5 | 20.8 | -75.2 | 6.2 | 2.8 | 1 | 0.097 |
| [time=1] | 0a | . | . | . | . | . | . |
| Age at T0 | -1.7 | 0.9 | -3.4 | 0.0 | 3.9 | 1 | 0.048 |
| [time=2] * Age at Time 0 | 1.1 | 0.8 | -0.4 | 2.7 | 2.1 | 1 | 0.152 |
| [time=1] * Age at Time 0 | 0a | . | . | . | . | . | . |
| Dependent Variable: P-PIRE-CDMI T0 100 | | | | | | | |

3) Gender

**P-PIRE-CDMI =gender (binary) + time + interaction (Gender*Time)**

| Parameter Estimates (n=35) | | | | | | | |
| --- | --- | --- | --- | --- | --- | --- | --- |
| Parameter | Estimate (ß) | Standard error | 95% Wald Confidence Interval | | Wald Chi-Square | df | Sig. |
|  |  |  | Lower | Upper |  |  |  |
| (Intercept) | 72.6 | 6.4 | 60.2 | 85.1 | 130.4 | 1 | 0 |
| [time=2] | -9.7 | 5.2 | -20.0 | 0.5 | 3.5 | 1 | 0.063 |
| [time=1] | 0a | . | . | . | . | . | . |
| [women] | -4.6 | 6.7 | -17.8 | 8.5 | 0.5 | 1 | 0.489 |
| [men] | 0a | . | . | . | . | . | . |
| [time=2] * [women] | 5.1 | 5.6 | -5.8 | 16.0 | 0.8 | 1 | 0.358 |
| [time=2] * [men] | 0a | . | . | . | . | . | . |
| [time=1] * [women] | 0a | . | . | . | . | . | . |
| [time=1] * [men] | 0a | . | . | . | . | . | . |
| Dependent Variable: P-PIRE-CDMI T0 100  N.B. “Did not specify group” not included because n=2 at T0 | | | | | | | |

4) Profession

**P-PIRE-CDMI =profession (binary) + time + interaction (Profession*Time)**

| Parameter Estimates (n=37) | | | | | | | |
| --- | --- | --- | --- | --- | --- | --- | --- |
| Parameter | Estimate (ß) | Standard error | 95% Wald Confidence Interval | | Wald Chi-Square | df | Sig. |
|  |  |  | Lower | Upper |  |  |  |
| (Intercept) | 72.7 | 2.5 | 67.7 | 77.6 | 826.6 | 1 | 0 |
| [time=2] | -8.1 | 2.5 | -13.0 | -3.1 | 10.3 | 1 | 0.001 |
| [time=1] | 0a | . | . | . | . | . | . |
| [OT] | -8.2 | 3.8 | -15.6 | -0.7 | 4.7 | 1 | 0.031 |
| [PT] | 0a | . | . | . | . | . | . |
| [time=2] * [OT] | 7.4 | 4.2 | -0.8 | 15.6 | 3.1 | 1 | 0.078 |
| [time=2] * [PT] | 0a | . | . | . | . | . | . |
| [time=1] * [OT] | 0a | . | . | . | . | . | . |
| [time=1] * [PT] | 0a | . | . | . | . | . | . |
| Dependent Variable: P-PIRE-CDMI T0 100 | | | | | | | |

5) Setting

**P-PIRE-CDMI =setting + time + interaction (setting*Time)**

| Parameter Estimates (n=37) | | | | | | | |
| --- | --- | --- | --- | --- | --- | --- | --- |
| Parameter | Estimate (ß) | Standard error | 95% Wald Confidence Interval | | Wald Chi-Square | df | Sig. |
|  |  |  | Lower | Upper |  |  |  |
| (Intercept) | 66.6 | 3.3 | 60.2 | 73.0 | 417.3 | 1.0 | 0 |
| [time=2] | -5.0 | 4.9 | -14.5 | 4.6 | 1.0 | 1.0 | 0.309 |
| [time=1] | 0a | . | . | . | . | . | . |
| [Missing or N‎/A setting] | 1.7 | 5.7 | -9.4 | 12.8 | 0.1 | 1.0 | 0.763 |
| [Private practice] | 2.0 | 4.8 | -7.4 | 11.3 | 0.2 | 1.0 | 0.677 |
| [Community agency, primary health care, home visiting agency, consulting firm] | -1.2 | 4.7 | -10.3 | 7.9 | 0.1 | 1.0 | 0.797 |
| [Long term care or rehabilitation center] | 19.1 | 3.3 | 12.8 | 25.5 | 34.5 | 1.0 | 0 |
| [Acute care] | 0a | . | . | . | . | . | . |
| [time=2] * [Missing or N‎/A setting] | -8.2 | 15.1 | -37.9 | 21.5 | 0.3 | 1.0 | 0.589 |
| [time=2] * [Private practice] | 0.9 | 6.1 | -11.1 | 12.8 | 0.0 | 1.0 | 0.886 |
| [time=2] * [Community agency, primary health care, home visiting agency, consulting firm] | 5.8 | 5.7 | -5.3 | 16.8 | 1.0 | 1.0 | 0.309 |
| [time=2] * [Long term care or rehabilitation center] | -9.3 | 4.9 | -18.9 | 0.2 | 3.7 | 1.0 | 0.055 |
| [time=2] * [Acute care] | 0a | . | . | . | . | . | . |
| [time=1] * [Missing or N‎/A ] | 0a |  |  |  |  |  |  |
| [time=1] * [Private practice] | 0a |  |  |  |  |  |  |
| [time=1] * [Community agency, primary health care, home visiting agency, consulting firm] | 0a |  |  |  |  |  |  |
| [time=1] * [Long term care or rehabilitation center] | 0a |  |  |  |  |  |  |
| [time=1] * [Acute care] | 0a |  |  |  |  |  |  |
| Dependent Variable: P-PIRE-CDMI T0 100  N.B. Missing or N/A group n=2 at T3; Long-term care or rehab center n=1 at T3 | | | | | | | |
